# The role of short-chain fatty acids on insulin sensitivity: a systematic review and meta-analysis

**DOI:** 10.1101/2022.11.22.22282645

**Authors:** Nhan H.T. Pham, Mugdha V. Joglekar, Wilson K.M. Wong, Najah T. Nassif, Ann M. Simpson, Anandwardhan A. Hardikar

## Abstract

**Context:** There is substantial evidence that reduced gut short chain fatty acids (SCFAs) are associated with obesity and type 2 diabetes, although findings from clinical interventions that could increase SCFAs are inconsistent.

**Objective:** This work aimed to assess the effect of SCFA intervention on fasting glucose, fasting insulin, and HOMA-IR by performing a systematic review and meta-analysis.

**Data sources:** Relevant published articles up to 28^th^ July 2022 were extracted from PubMed and EMBASE using the medical subject heading (MeSH) of the defined keywords [(short-chain fatty acids AND (obesity OR diabetes OR insulin sensitivity)] and their synonyms. Data analyses were performed using recommended Cochrane meta-analysis checklist and PRISMA guidelines and were registered in the PROSPERO system (ID: CRD42021257248).

**Data extraction:** Clinical studies and trials that measured SCFAs along with reporting glucose homeostasis parameters were included in the analysis. Standardized mean differences (SMD) with 95% confidence intervals (CI) were calculated using a random-effects model in the data extraction tool Review Manager version 5.4 (RevMan 5.4). The risk of bias assessment was performed following the Cochrane checklist for randomised and crossover studies.

**Data analysis:** We identified 6,040 non-duplicate studies; 23 met defined criteria and reported fasting insulin, fasting glucose, or HOMA-IR as well as a measured post-intervention change in SCFA concentrations. Our meta-analyses indicated that fasting insulin concentrations significantly reduced (overall effect: SMD = -0.15; 95% CI = -0.29, -0.01, p = 0.04) between placebo and treatment groups at the end of the intervention. Post-intervention increase in SCFAs had an even significant effect on lowering fasting insulin (p=0.008). Elevated levels of SCFAs were also associated with beneficial effects on HOMA-IR (p < 0.00001) compared to their baseline. Fasting glucose concentrations did not show any significant change.

**Conclusions:** Increased post-intervention SCFA is associated with lower fasting insulin, offering a beneficial effect on insulin sensitivity.

## Introduction

Type 2 diabetes mellitus (T2D) is characterised by a reduction in β-cell function as well as insulin resistance, wherein skeletal muscle, liver, and adipocytes cannot uptake sufficient amounts of glucose ^1^. T2D leads to life-threatening complications such as neuropathy, retinopathy, and cardiovascular diseases. According to the World Health Organization, there are over 537 million adults living with T2D and this number is estimated to reach 783 million by 2045 ^2^.

Along with T2D, overweight (body mass index/BMI of 25 to <30) and obesity (BMI ≥ 30) are considered major health problems, currently affecting approximately two billion people worldwide ^3,4^. Obesity is a major health risk for the development of cardiovascular disease, hypertension, and stroke. It is also known to dramatically increase the risk of T2D, wherein 90% of individuals with T2D are obese or overweight ^5^. Beneficial effects of gut microbiota and short-chain fatty acids (SCFAs) on metabolic health in obesity ^6,7^ and T2D ^8,9^, have been reported. The diversity in the gut microbiome is well recognised between lean and obese individuals ^6,7^ as well as between individuals with and without diabetes ^10,11^. Gut microbes produce SCFAs, mainly acetate, propionate, and butyrate, through the fermentation of resistant starch or soluble fibre. These molecules are shown to induce the expression of gut hormones such as peptide tyrosine-tyrosine (PYY) and glucagon-like peptide 1 (GLP-1) via free fatty acid receptors (G-Protein Coupled Receptor, GPR 41/43) ^12,13^, leading to appetite suppression ^14^ and improvement of glucose tolerance and insulin sensitivity ^15,16^. In addition, it has been shown that fecal transplantation from lean donors to obese recipients altered their gut microbial composition and improved insulin sensitivity ^17,18^.

SCFAs can be administered directly, as sodium salts or esters, or indirectly as pre-/probiotics or high-fibre diets. Multiple clinical trials have aimed at improving SCFAs in participants with impaired insulin sensitivity, pre-existing overweight/obesity, and multiple gastrointestinal abnormalities. Many meta-analyses are available on the topic of SCFAs in human clinical trials, however, most of these focus on irritable bowel disease ^19-21^, inflammatory markers ^22,23^, obesity ^24,25^, or gut biota ^26,27^. Relevant previous meta-analyses (**Table S1**) report effects on insulin and glucose concentrations after consumption of specific diet such as fibre, resistant starch or wholegrains ^28-33^ or synbiotics ^34^ and probiotics ^35,36^. Changes in total SCFAs were analysed only in one review along with *Bifidobacteria* and fasting glucose ^33^; however this study did not examine the effect of SCFA changes on fasting insulin or HOMA-IR. To our knowledge, there is no systematic review and meta-analyses that assessed the effects of post-intervention changes in SCFAs on insulin sensitivity in humans. To address this research gap, we analysed clinical studies (randomised clinical trials, observational, or treatment-only studies) that reported actual values of either glucose, insulin, or HOMA-IR as well as measured SCFAs following interventions. Our systematic review and meta-analysis are inclusive of different interventions that have the potential to alter SCFAs and are focussed on studies that confirm a change in SCFA concentrations post-intervention.

## Methods

### Data sources and search

We carried out a systematic search using PubMed and EMBASE via Ovid for studies published from January 01, 1959, to July 28, 2022. There were no restrictions on the use of Medical Subject Heading (MeSH) terms and their synonyms. The keywords used for the literature search were “short-chain fatty acids AND (obesity OR diabetes OR insulin sensitivity)”. All studies were screened based on their title, abstract, and full-text (**Figure 1**), following the recommended Cochrane meta-analysis checklist and PRISMA guidelines ^37^. Search strategy, study eligibility, data extraction, and analysis of this systematic review were pre-registered in the PROSPERO system (ID: CRD42021257248).

**Figure 1:**
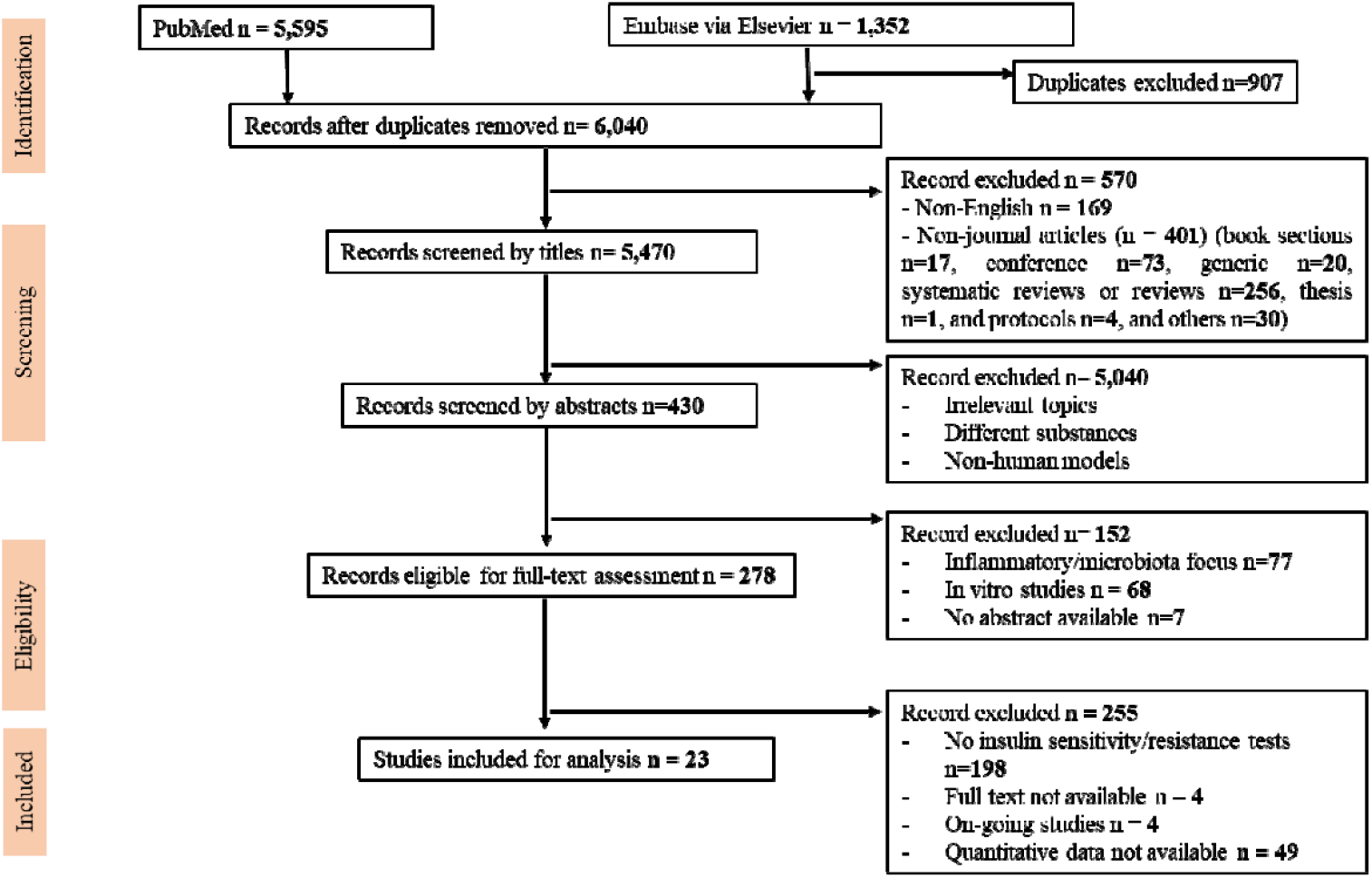
Study flow chart (PRISMA) diagram summarizing the process for the selection of papers for the analysis of the effects of SCFAs on fasting insulin, fasting glucose and HOMA-IR.

### Inclusion and Exclusion criteria

Papers were included for screening if they contained the keywords (as defined in the literature search section above) and PICOS criteria summarised in **Table 1**. Exclusion criteria were defined as (1) not human studies such as animal studies or cell culture studies, (2) not published in the English language, and (3) publication types other than peer-reviewed original papers, such as systematic reviews, meta-analysis, reviews, conference publications, non-article papers, generic writing, thesis chapters, and case reports, (4) not providing data for fasting glucose, fasting insulin or HOMA-IR, and (5) not reporting SCFA concentrations pre- and post-intervention.

**Table 1:** PICOS criteria for inclusion and exclusion of studies analysed in this systematic review and meta-analysis. PICOS stands for participants, intervention, comparison, outcomes, and study design.

|  | **Inclusion** | **Exclusion** |
| --- | --- | --- |
| **Participants** | Healthy adults, obese/overweight adults, or adults with underlying conditions | No restriction on the human participants. Animal and *in vitro* cellular studies were excluded. |
| **Interventions** | Direct SCFA interventions (oral/IV infusion/enema of sodium acetate, propionate, or butyrate alone or mixed) or dietary supplementary (resistant starch, inulin, or chemically modified fibres) | Direct supplementation of substances other than three SCFAs (acetate, propionate or butyrate) |
| **Comparators** | Placebo vs intervention (endpoint); baseline vs endpoint (intervention group) | Two different treatments or doses or intermediate time points |
| **Outcomes** | The actual concentrations of fasting insulin, fasting glucose, or HOMA-IR as well as reporting of (actual concentration/bar graphs/in text) change in plasma or faecal levels of SCFAs at the end of intervention | Studies not reporting the concentrations of fasting insulin, fasting glucose, or HOMA-IR (such as AUCs, regression/correlation coefficients, bar graphs, difference in concentrations), AND not providing change in SCFAs |
| **Study designs** | Randomized Clinical Trials, crossover studies or treatment-only studies | Non-original papers (reviews, systematic reviews, conference reports, clinical trial protocols, ongoing studies) and Non-English papers |

### Data extraction

Results from the search were added to an EndNote X8 ^38^ (Clarivate Analytics, Toronto, Canada) library and in an MS-Excel worksheet for data screening and extraction (**Figure 1)**. The first step of removing duplicate hits from the two databases EMBASE and PubMed was performed in the EndNote library, where papers with identical titles and authors were marked as duplicates and removed. In the screening step, titles and abstracts were assessed for eligibility, and articles were excluded if they met any of the exclusion criteria. The 23 qualifying papers were then reviewed in full-text versions and included in the meta-analysis that were 1) original human clinical studies, 2) provided values (not AUCs or coefficients) of fasting glucose, insulin, and/or insulin resistance score/HOMA-IR, and 3) verified plasma/faecal SCFAs before and/or after the intervention. All values were converted to mean ± SD, mmol/L for glucose, and μU/mL for insulin. Reported units for glucose and insulin were made consistent ^39^. In two ^40 41^ of the 23 studies, units or standard deviation values were confirmed with the authors of the original articles via email correspondence. Sub-group analyses were performed to investigate differences in fasting insulin, glucose, or HOMA-IR between placebo and treatment groups, or between the baseline and the endpoint. Two researchers (NHTP and MVJ) performed the search and data extraction independently and then cross-checked.

### Quality of Evidence and Risk of Bias

The assessments of risk of bias and quality of evidence were conducted following the Cochrane recommendations ^42^, and outcomes were presented using RevMan (Version 5.4, The Cochrane Collaboration, 2020) ^43^. The criteria included random sequence generation (selection bias), allocation concealment (selection bias), blinding of participants and personnel (performance bias), blinding of outcome assessment (detection bias), incomplete outcome data (attrition bias), selective bias (reporting bias), and other bias. Each criterion was assigned as either high risk, low risk, or unclear. Treatment-only studies were not included in the selection, performance, and detection bias as all participants and associated clinicians were aware of treatment allocation. Several of the studies had a crossover design. Therefore, we also assessed three additional criteria for crossover studies including appropriate crossover design, carry-over effects, and unbiased data (reporting data at all stages of the trial) ^44^.

### Data Synthesis and Statistical analysis

Data synthesis and statistical analysis were carried out using Microsoft Excel (MS-Excel ver. 2016; Microsoft Corporation, Redmond, WA, USA) and RevMan 5.4. MS-Excel was used to tabulate and/or summarize the data collected, for calculating insulin and glucose values at appropriate units as well as to plot stacked bar graphs. RevMan 5.4 software was used to produce forest plots. All data were entered as mean± SD. Standardized mean differences (SMD) with 95% confidence intervals (CI) using the random effects model and inverse variance statistical method were performed between the placebo group and the intervention group, or between the baseline and the endpoint. We also carried out the same analysis using the fixed effects model to ensure the robustness of our calculations. All the analyses were performed in RevMan 5.4. A P-value < 0.05 was considered as significant. SMD values for SCFAs, insulin, and glucose were calculated in RevMan 5.4 software and used for Spearman correlation analysis. Correlation matrix was generated using corrplot (0.92), Hmisc (4.6.0), dplyr (1.0.7) packages and R (version 3.6.1) in Rstudio (2021.09.0, Build 351, R Foundation for Statistical Computing, Vienna, Austria). The correlation significance was analysed and verified on GraphPad Prism version 9.4.1 (GraphPad Software, San Diego, CA, USA).

## Results

### Characteristics of studies included for meta-analysis

The PRISMA flow diagram for the stepwise selection of studies is shown in **Figure 1**. The initial search using the defined MeSH terms in PubMed and EMBASE via Ovid identified 6,040 papers (after excluding 907 duplicates). In the screening step, 570 articles were removed as they were non-English papers or non-journal articles. Following title and abstract screening on the remaining 5,470 papers, 278 papers were deemed eligible for full-text assessment. A majority (198 studies) of these studies did not perform insulin sensitivity/resistance tests while 49 papers did not present quantitative data on fasting insulin or fasting glucose and were therefore removed from the meta-analysis. Twenty-three studies were included in the final selection (**Table 2**) that contained actual values for fasting glucose and insulin, and therefore included in the meta-analysis ^40,41,45-65^.

**Table 2:**
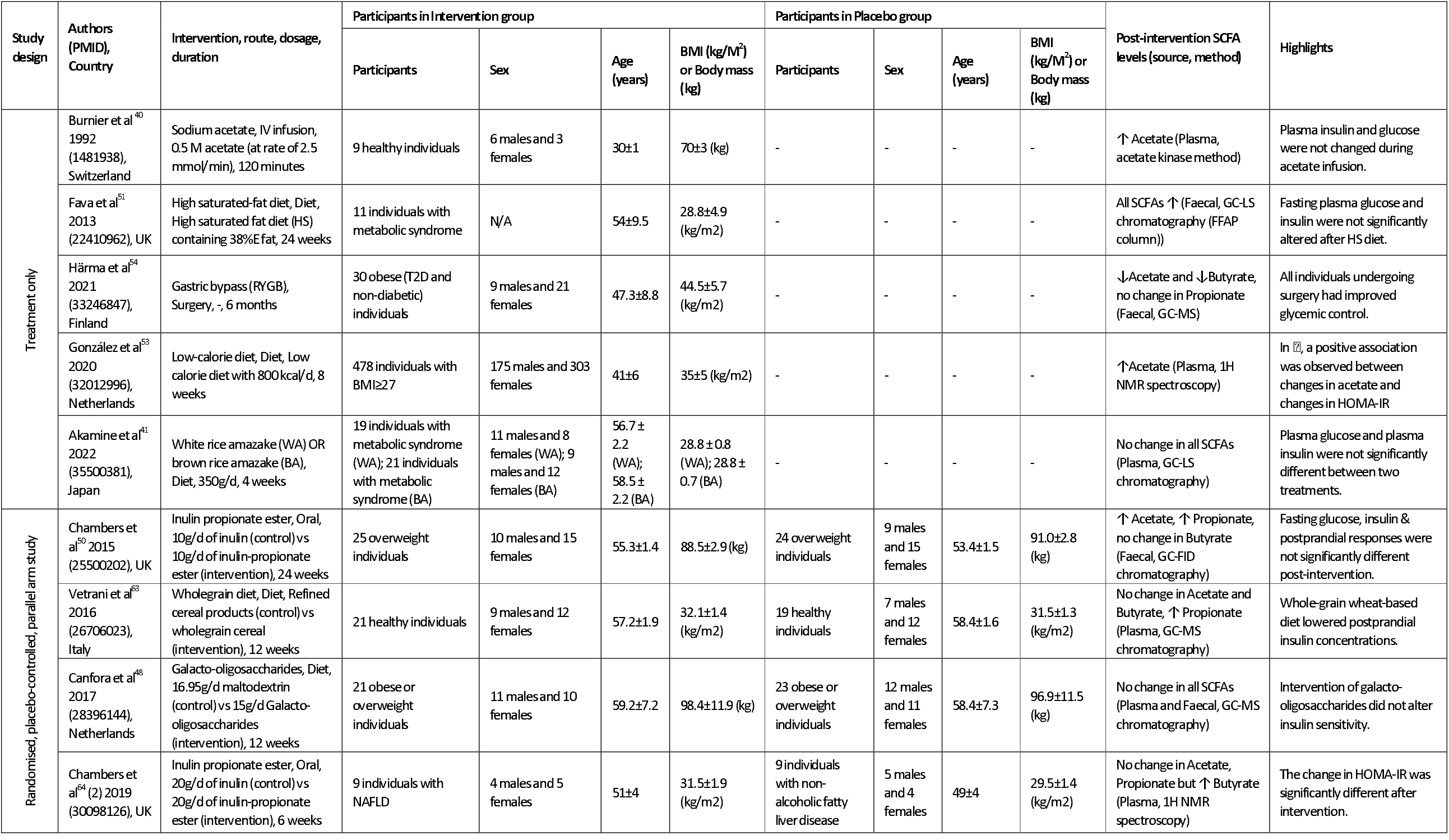
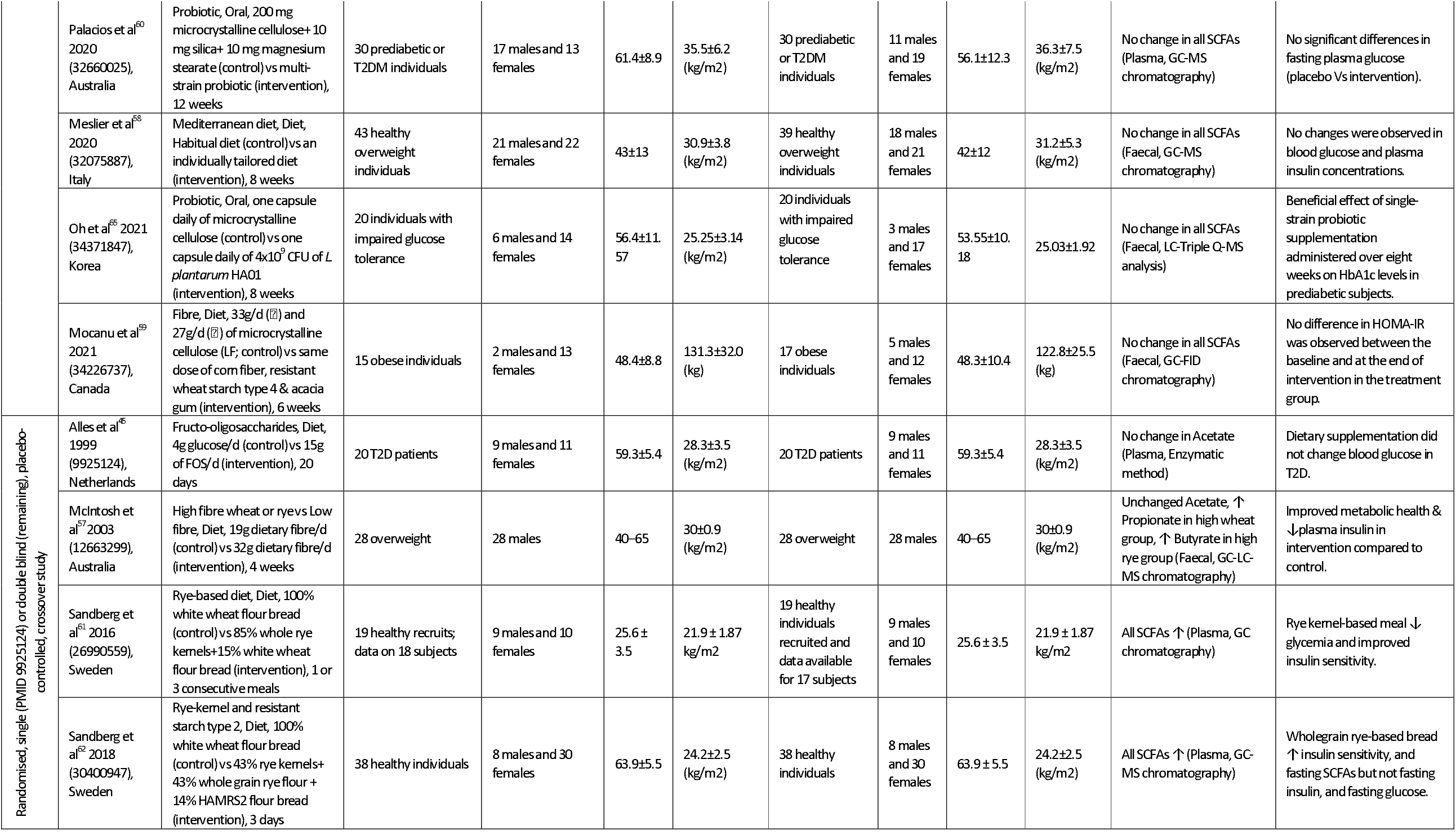
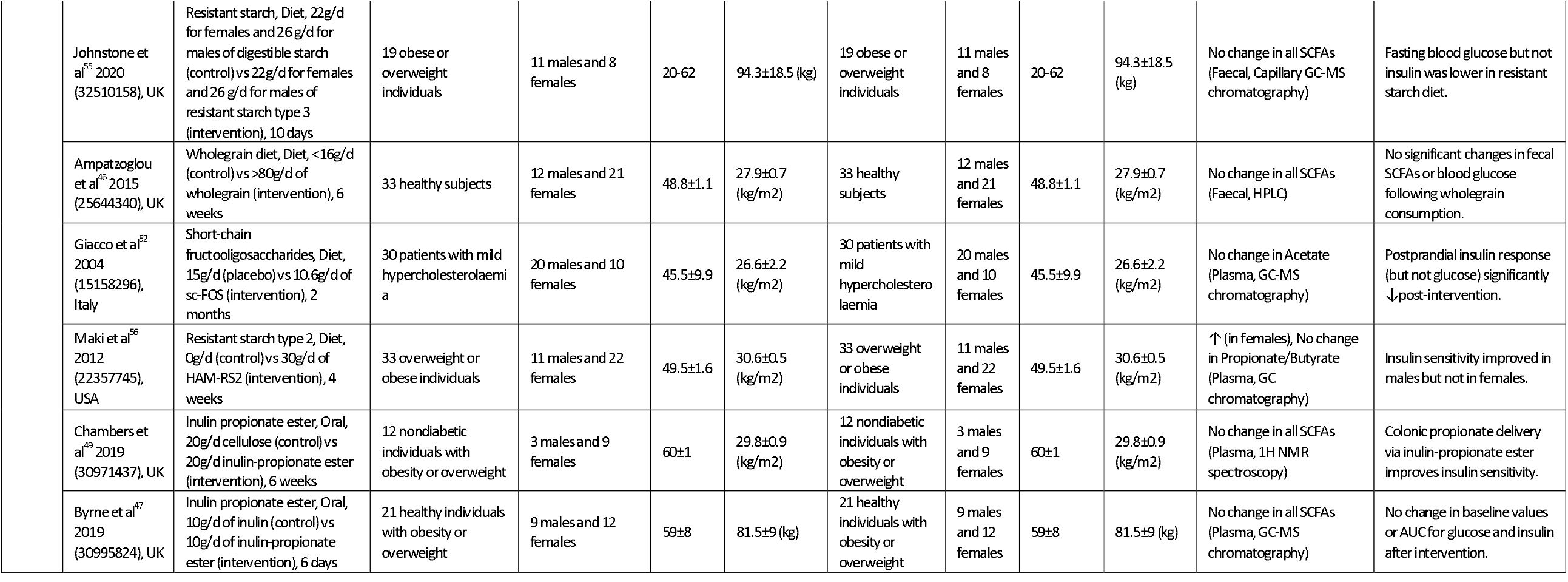
Summary of studies included in the meta-analysis, which details study design, intervention, duration, dose, participant information as well as changes and source of SCFAs. The age, weight or BMI values are reported as per the original article. The effects of the intervention on glucose-insulin parameters are highlighted.

**Table 2** summarises the 23 studies used in the meta-analysis. Analyses include data on a total of 1,186 study participants from all categories (healthy 12%, obese/overweight 67%, and people with underlying conditions including diabetes 21%). Out of the 23 studies, 17 had dietary interventions, four studies provided propionate as inulin propionate ester, and one study had gastric bypass treatment, while the other study had sodium acetate infusion. Most of the studies followed intervention for as little as 10 days and up to six months, while only four studies had interventions for less than one-week duration. Either one or all the three major SCFAs (acetate, propionate, and butyrate), from plasma or faeces, were measured in all studies using chromatography, spectroscopy, or enzymatic methods. Although these 23 papers measure plasma/faecal SCFAs, few of these do not report the actual SCFA concentrations. It was therefore difficult to perform a meta-analysis for SCFA concentrations in different interventions. We, therefore, divided our analysis into two subgroups (one with increased SCFAs and the other without increased SCFA at the end of the intervention; **Figures 2 to 4**).

**Figure 2:**
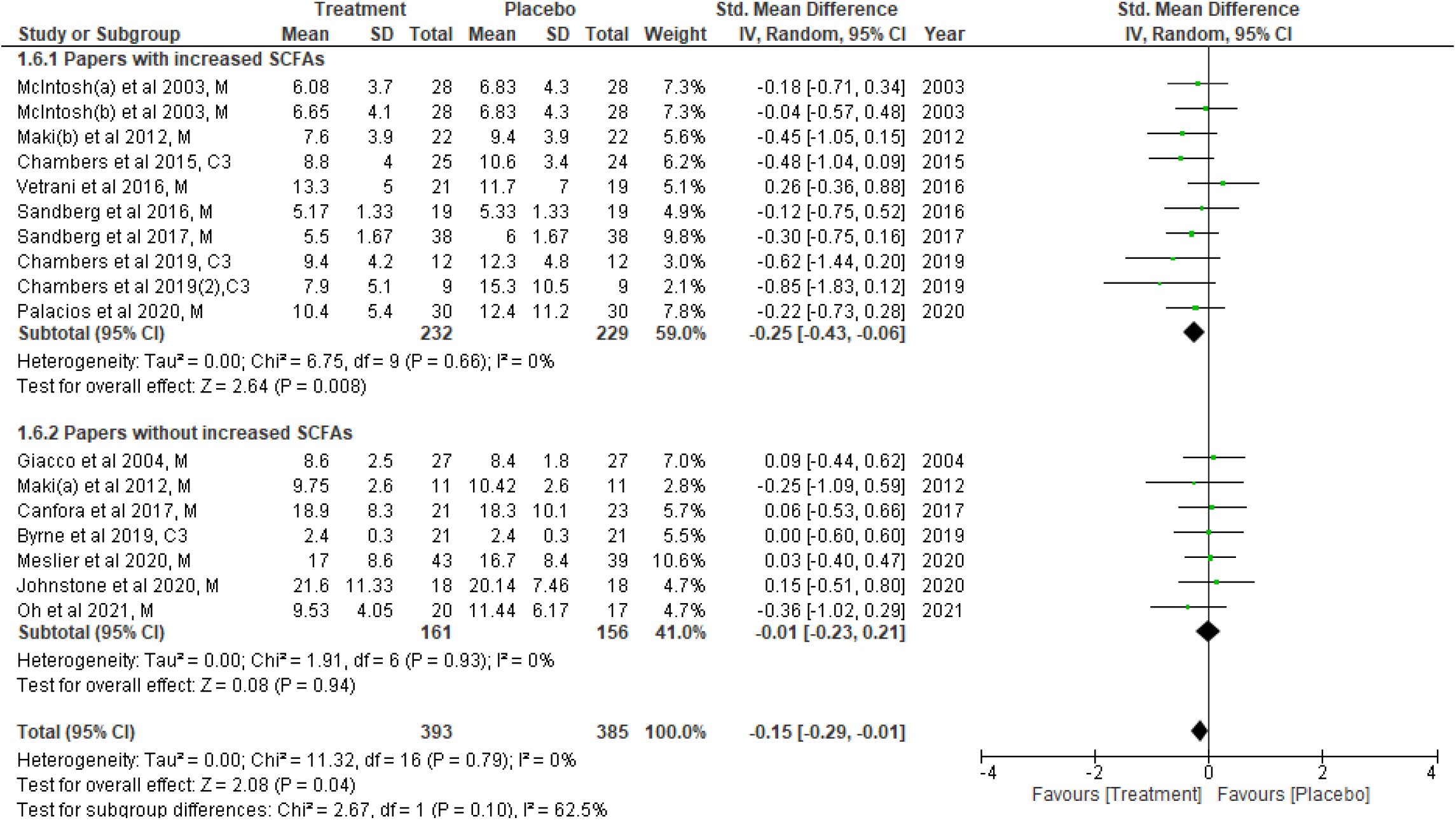
Fasting insulin concentrations from each study were compared between placebo and treatment at the endpoint (end of intervention period). Data were presented as SMD of fasting insulin (μU/mL) and were separated into two subgroups: one with evidence of an increase in SCFA concentration(s) post-intervention, and the other without any change in SCFAs post-intervention. The type of intervention (M: meal/mixed and C3: propionate) is noted at the end of each study. df: degrees of freedom, IV: inverse variance, CI: confidence interval.

### Changes in glucose homeostasis parameters between placebo and intervention groups at the endpoint

The effects of SCFAs on fasting insulin (**Figure 2**), fasting glucose (**Figure 3**), and HOMA-IR (**Figure 4**), are stratified for studies (from **Table 2**) that confirm a significant increase or no change in SCFA concentrations post-intervention. In a comparison between placebo and intervention groups at the endpoint (final assessment after intervention), fasting insulin was significantly lower (SMD = -0.25; 95% CI = -0.43, -0.06; p = 0.008) in the treatment group when SCFA concentrations were confirmed to have increased at the end of intervention (**Figure 2**). The overall effect on fasting insulin concentrations was also significant (SMD = -0.15; 95% CI = -0.29, -0.01; p = 0.04; **Figure 2**). In contrast, fasting glucose (SMD = -0.06; 95% CI = -0.24, 0.13, p = 0.55; **Figure 3**) and HOMA-IR (SMD = -0.22; 95% CI = -0.48, 0.05, p = 0.11; **Figure 4**) did not differ significantly between treatment and control arms for studies wherein SCFA concentrations increased post-intervention. For the subgroup of studies without increase in SCFAs, there were no significant differences in fasting insulin, fasting glucose, and HOMA-IR between the control and treatment arms (**Figures 2 to 4**).

**Figure 3:**
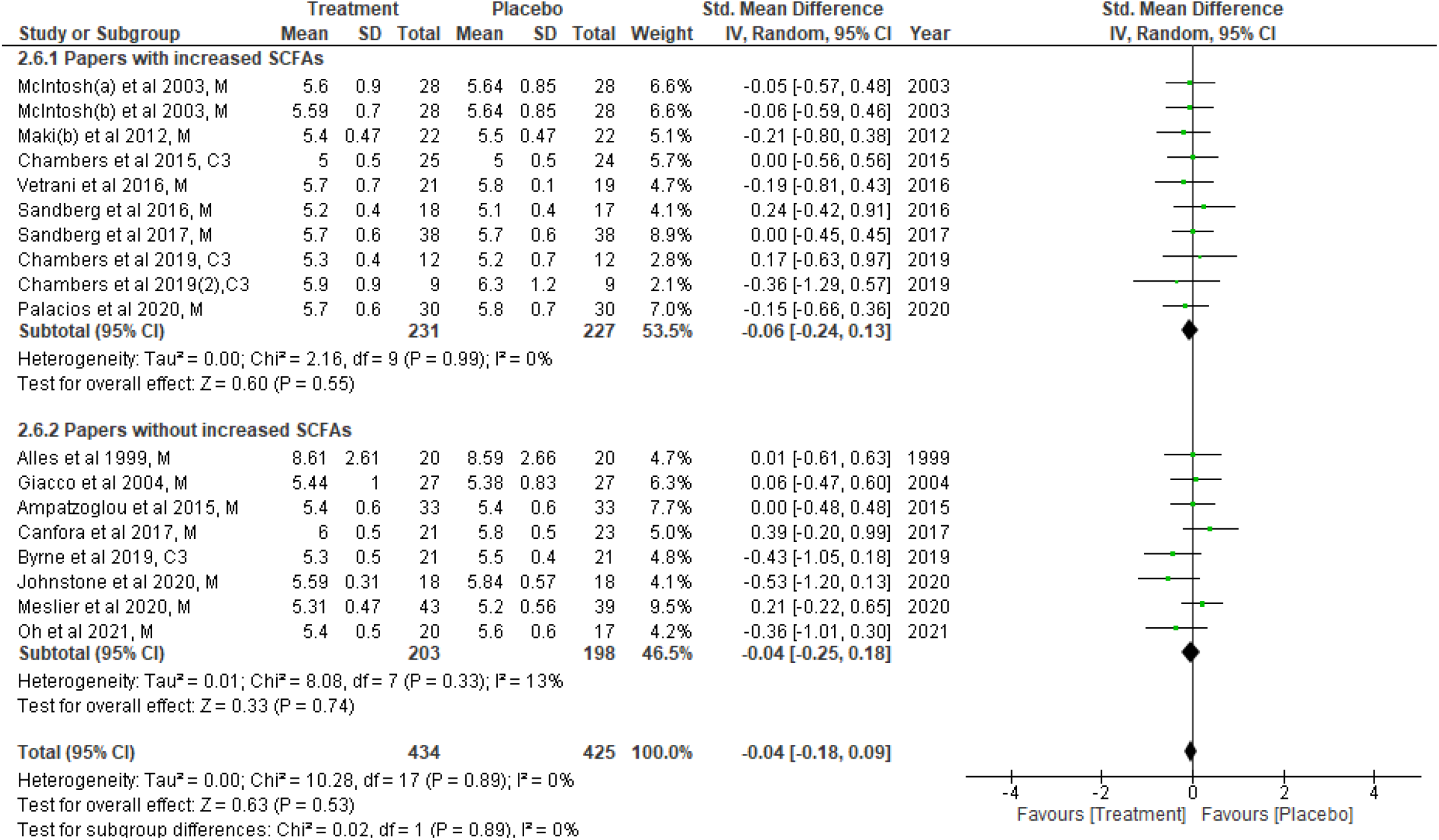
Fasting glucose concentrations from each study were compared between placebo vs treatment at the endpoint (end of intervention period). Data were presented as SMD of fasting glucose (mmol/L) and were separated into two subgroups: one with evidence of an increase in SCFA concentration(s) post-intervention, and the other without any change in SCFAs post-intervention. The type of intervention (M: meal/mixed and C3: propionate) is noted at the end of each study. df: degrees of freedom, IV: inverse variance, CI: confidence interval.

**Figure 4:**
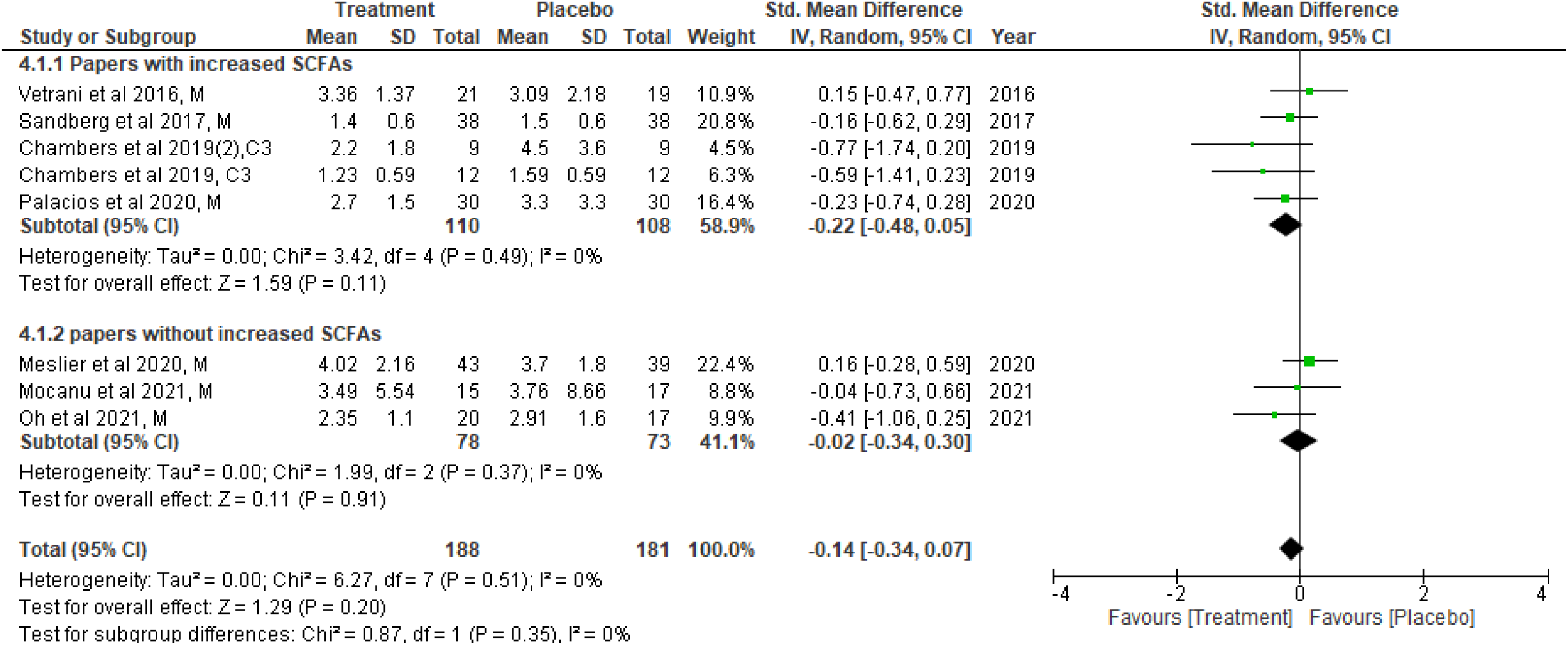
HOMA-IR values were compared between the placebo and treatment at the endpoint (end of intervention period). Data were presented as SMD of HOMA-IR and separated into two subgroups: one with evidence of an increase in SCFA concentration(s) post-intervention, and the other without any change in SCFAs post-intervention. Type of intervention (M: meal/mixed and C3: propionate) is noted at the end of each study. df: degrees of freedom, IV: inverse variance, CI: confidence interval.

When fasting insulin values were analysed based on direct SCFA or indirect dietary treatment, a significant reduction of fasting insulin was observed only after direct intervention (SMD = -0.39; 95% CI = -0.74, -0.05; p = 0.03) but not after the indirect (dietary) intervention compared to their controls (**Figure S1**). Further subgrouping into direct intervention with evidence of increased SCFAs showed significant reduction of fasting insulin (SMD = -0.58; 95% CI = -1.01, -0.16; p = 0.007) (**Figure S2**). However, it should be noted that all the three studies included in this analysis were from the same group and for inulin-propionate ester intervention ^49,50,64^. Similar sub analyses for the source of SCFA measurement (faecal or plasma) revealed that elevated plasma SCFAs were associated with a significant reduction in fasting insulin concentrations (SMD= -0.22, 95% CI = -0.43, -0.01, p= 0.04: **Figure S3**). Since the beneficial effects on glycemic parameters were seen when SCFAs were higher, we performed a correlation analysis on the changes in SCFA, insulin, and glucose concentrations between placebo and intervention at the endpoint (**Figure S4**). Out of the 23 studies, only 8 studies could be included for this analysis as only those reported the actual values of SCFA concentrations (**Table S2**). Notably, we found that the standardised mean difference (SMD) of acetate between the placebo and intervention group was significantly and inversely correlated with that of fasting insulin (r=-0.64, p=0.048).

### Improvement in glucose-insulin measurements relative to study baseline

We also analysed changes in fasting insulin, glucose as well as HOMA-IR, with reference to their baseline measurements in the intervention group and the placebo group. A significantly lower fasting insulin was observed at the end of intervention (**Figure 5**) compared to baseline (SMD = -0.22; 95% CI = -0.43, -0.01; p = 0.04) in studies that confirm increased SCFA concentrations. The overall effect on fasting insulin was also significant when all studies were included (SMD=-0.18, 95% CI = -0.34, -0.03, p=0.02: **Figure 5**). Fasting glucose did not demonstrate any significant changes relative to baseline in the meta-analysis (**Figure 6**). On the other hand, HOMA-IR was significantly different (SMD = -0.44; 95% CI = -0.56, -0.32; p < 0.00001) in studies with increased post-intervention SCFA concentrations. This significance was also observed when all the studies (demonstrating increased/no-change/decreased SCFAs post-intervention) were included in the analysis (SMD = -0.37; 95% CI = -0.48, -0.27; p <0.00001) (**Figure 7**). In our analysis for HOMA-IR, one study ^53^, had more participants than others. Baseline versus endpoint comparison in the placebo group alone did not demonstrate any significant improvement in fasting insulin, fasting glucose, and HOMA-IR (**Figure S5, S6 and S7**) respectively.

**Figure 5:**
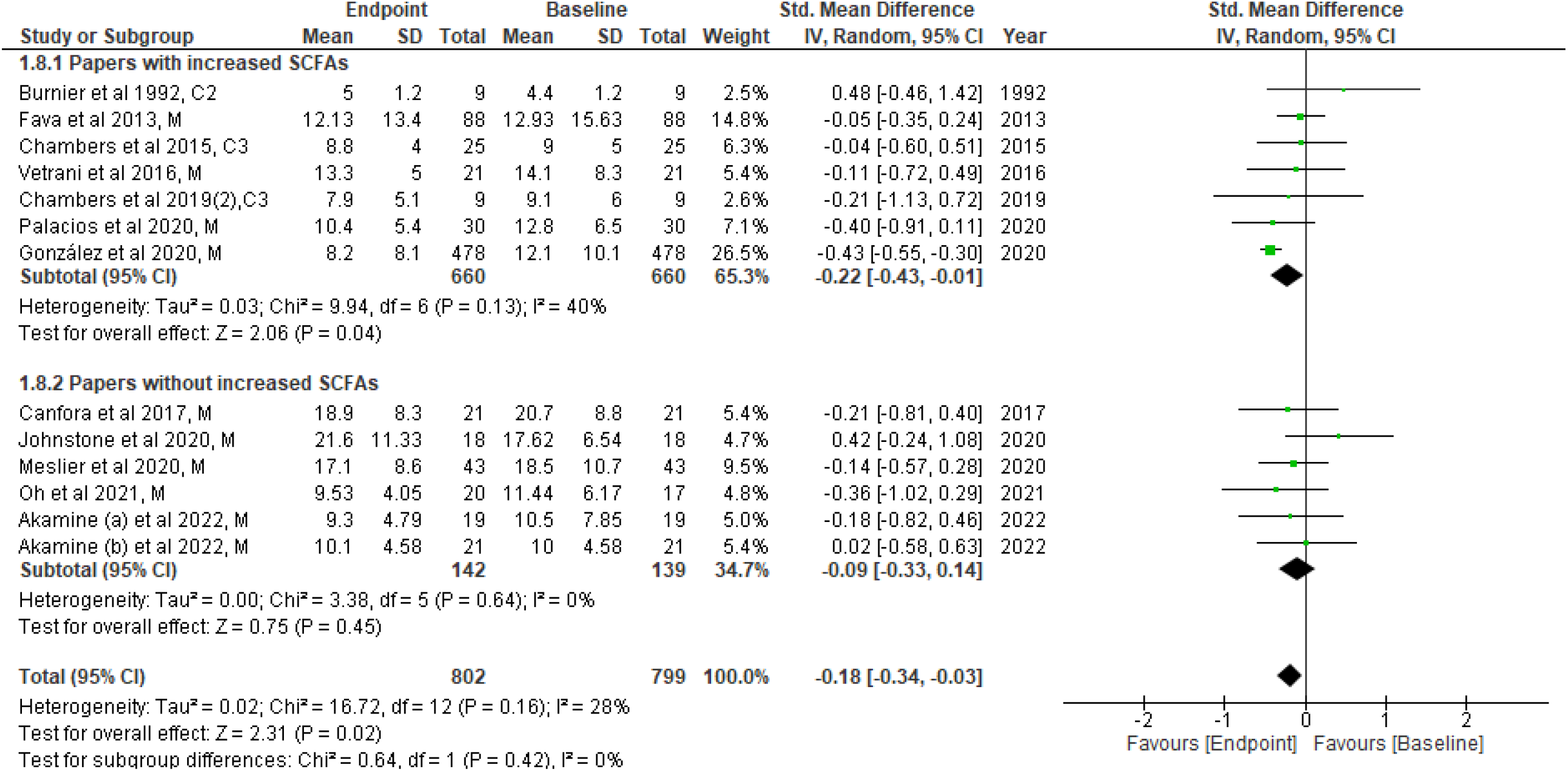
Fasting insulin concentrations at baseline and endpoint (end of intervention) from each study were compared in the treatment group. Data were presented as the SMD of fasting insulin (μU/mL) and were separated into two subgroups: one with evidence of an increase in SCFA concentration(s) post-intervention, and the other without any change in SCFAs post-intervention. Type of intervention (M: meal/mixed, C3: propionate and C2: acetate) is noted at the end of each study. df: degrees of freedom, IV: inverse variance, CI: confidence interval.

**Figure 6:**
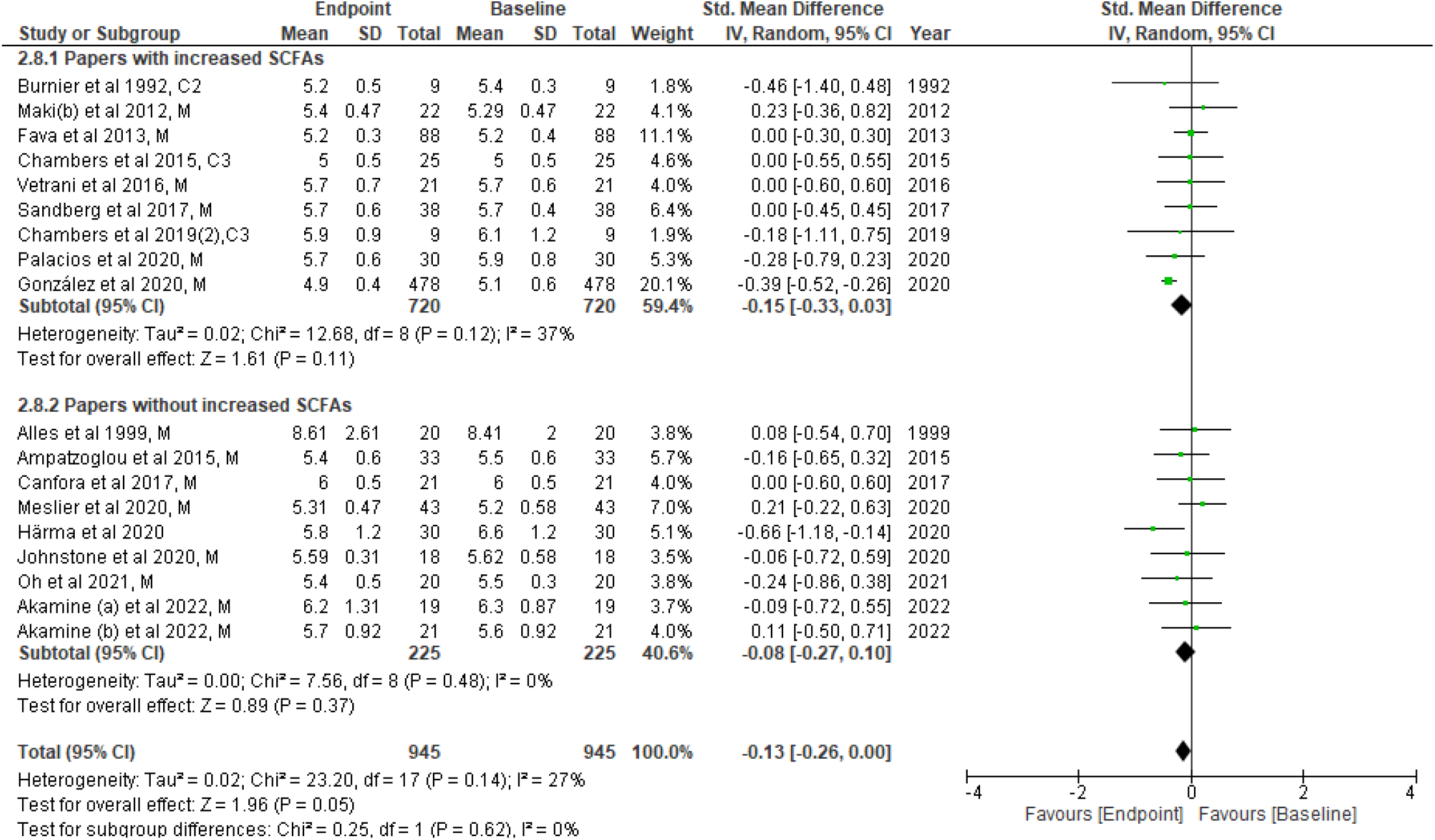
Fasting glucose concentrations at baseline and endpoint (end of intervention) from each study were compared in the treatment group. Data were presented as SMD of fasting insulin (mmol/L) and were separated into two subgroups: one with evidence of an increase in SCFA concentration(s) post-intervention, and the other without any change in SCFAs post-intervention. Type of intervention (M: meal/mixed, C3: propionate and C2: acetate) is noted at the end of each study. df: degrees of freedom, IV: inverse variance, CI: confidence interval.

**Figure 7:**
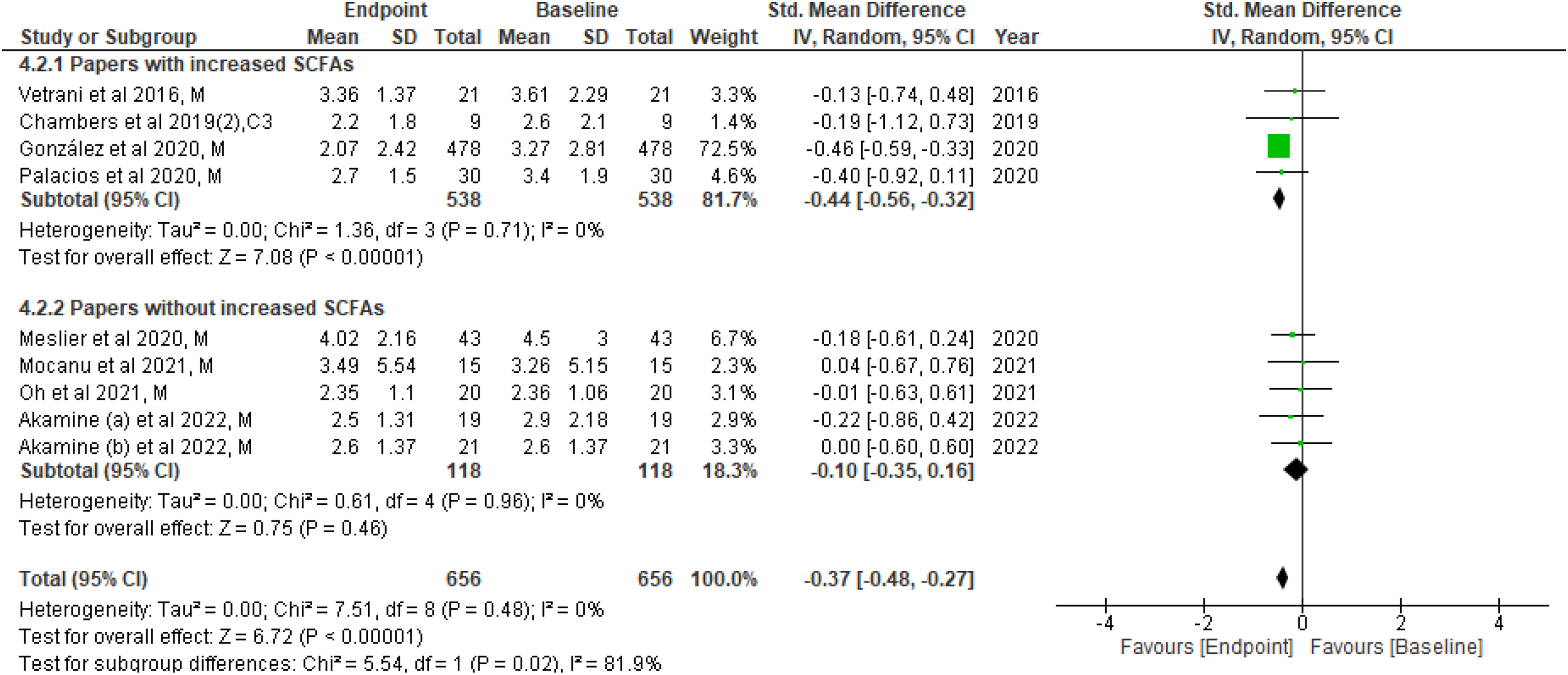
HOMA-IR values at baseline and endpoint (end of intervention) from available studies were compared in the treatment group. Data were presented as SMD of HOMA-IR and were separated into two subgroups: one with evidence of an increase in SCFA concentration(s) post-intervention, and the other without any change in SCFAs post-intervention. Type of intervention (M: meal/mixed and C3: propionate) is noted at the end of each study. df: degrees of freedom, IV: inverse variance, CI: confidence interval.

### Risk of bias assessment

The quality assessment of the 23 papers included in the meta-analysis is presented in **Figure S8**. As mentioned in the methods section, crossover studies were analysed separately ^45-47,49,52,55-57,61,62^ (**Figure S8A**). Several of the studies did not report sufficient details of unbiased data presentation or carry-over effects to assign low or high risks, therefore, they had to be classified as unclear risks. Allocation concealment criteria was not described in 80% of the papers and is the highest factor with unclear risks.

Similarly, unbiased data (availability of study results at every time point during crossover) reporting was not clear in most of the studies. A high risk of bias was largely found for blinding criteria. Since most of the studies are dietary interventions, blinding may not be always applicable (i.e. the participants and the researchers could see what foods were given). In the parallel-arm trials, the risk of bias was much lower in most of the studies (**Figure S8B**). Five treatment-only studies ^40,41,51,53,54^ were not included for selection, performance, and detection bias analyses, since all participants received the same treatment, and any selection/blinding was not applicable. These studies are denoted by the white area in the graphs (**Figure S8B**).

## Discussion

This systematic review included 23 papers in the meta-analysis consisting of a varied group of participants. The main finding was significantly lower fasting insulin in the intervention group that received SCFAs directly, as well as in the group that had evidence of an increase in SCFAs following intervention compared to the placebo group. When treatment baseline and endpoint values were compared, fasting insulin and HOMA-IR were significantly improved at the endpoint, which was not observed for the placebo baseline vs endpoint comparison. Similar results were achieved when using the fixed effects model, thus confirming the robustness of this study.

The underlying mechanism linking SCFAs to T2D has not been fully elucidated. A mechanism involving gut microbes/microbial metabolites (eg. colonic SCFAs) regulating incretin hormone expression, insulin secretion, and glucose homeostasis is likely and needs validation. Incretin hormones such as glucose-dependent insulinotropic polypeptide (GIP) and glucagon-like peptide 1 (GLP-1) are known to regulate insulin secretion following oral glucose intake ^66^. GLP-1 also plays a significant role in improving glucose tolerance and insulin sensitivity ^15,16^. Interestingly, SCFAs lowered fasting insulin and improved HOMA-IR, without affecting fasting glucose. It would have been informative to understand the effects of the increase in SCFAs on GLP-1 concentrations, however only six studies performed GLP-1 measurements ^47-49,58,61,62^, and only three of these studies mentioned the use of the necessary dipeptidyl peptidase 4 (DPP-4) inhibitors during the collection of samples ^48,61,62^. Meta-analysis to understand the effect of changes in SCFAs on GLP-1 was inconclusive in this same set of papers.

Sodium acetate was the first SCFA to be applied in human studies in the 1980s via gastric or IV infusion with various dosages ^67-70^. Notably, most of the studies during the last decade have focused on the use of sodium propionate, sodium butyrate, or SCFA mixtures via the oral route for extended periods of time (up to 6 months), with dosages ranging from 1-10 g per day. These studies, and the long periods of SCFA intervention underscores their safety in human clinical studies/trials. To date, there have been no reports of any adverse side effects of SCFAs on human health. A very recent systematic review and meta-analysis assessed the effects of acute and chronic administration of SCFAs and vinegar on glycemic control ^71^. Interestingly, they did not observe a significant effect of acute acetate and propionate as well as chronic propionate treatment on glucose and insulin concentrations. However, this study did not stratify their analyses based on changes in SCFA concentrations post-intervention. Based on our quantitative analysis, only those studies with evidence of increase in SCFA concentrations had significantly lower fasting insulin, whereas studies without increase in SCFAs did not change insulin concentrations.

SCFAs are typically measured in faecal or plasma samples. In faeces, they are present in larger quantities^72^ and faecal SCFAs are considered as an indirect representation of SCFA production in the colon. SCFAs that are present in the circulation are in smaller quantities and usually reflect the amount that is remaining after their uptake in colonocytes and then in the liver^73^. Nonetheless, circulating SCFAs are shown to be better associated with metabolic health than faecal SCFAs ^74^. Our sub-analysis also confirmed significant effect on fasting insulin concentrations when circulating SCFAs are increased. One of the potential reasons for higher variability in faecal SCFAs could be the sample collection, storage, and processing methods ^75^. Standardisation of approaches in SCFA measurements are urgently needed.

Our meta-analysis identified that direct administration of SCFAs is beneficial than indirect methods including varied diets. It is notable that the use of SCFAs via oral administration is not favourable due to their unpleasant taste and odour^76^. Recent studies have been developing novel targeted delivery systems that can deliver SCFAs into the colons or the circulating system ^77^. In our analysis, there was only one study that had a direct infusion of SCFAs (i.e., acetate infusion) ^40^ and reported increases in circulating (plasma/serum) SCFAs. All the remaining studies that provided SCFAs as either capsules or powders or meals reported variable SCFA concentrations ^47,49,50,64^ at the endpoint. Apart from potential differences in faecal and circulating SCFA concentrations, another reason for this variation could be a different method of compliance assessment. Three trials ^49,50,64^ maintained regular communication between participants and investigators to encourage high compliance, and the participants returned their used and unused sachets to calculate the compliance rate at the end of the study. Future studies should ensure that compliance with intervention is assessed and reported.

Due to the invasive nature of infusion studies, the direct infusion of acetate was performed for a short period of time ^40^. However, studies that confirmed an increase in SCFA concentrations, were the ones that reported a lowering effect on fasting insulin. Therefore, it would be interesting to understand the long-term effects of SCFA infusion on insulin sensitivity. Clinical trials addressing this question are warranted. Interestingly, a separate systematic review and meta-analysis confirmed that the left colon may contribute to the maintenance of glucose homeostasis ^78^. Since human GLP-1-producing L-cells are found at the highest density in the left colon ^79^, removal of the left colon is, not surprisingly, associated with T2D. SCFAs provide a unique opportunity to manipulate the expression of incretin hormone expression and more studies in this area are needed.

### Limitations

Limitations of this meta-analysis are mainly due to the heterogeneity, which was unavoidable due to the nature of our study question on SCFAs and insulin sensitivity. The factors contributing to the high heterogeneity were age, sex, sample size, diet, lifestyle, background health conditions, types of SCFAs measured (acetate/propionate/butyrate/all) as well as a source of SCFAs (faeces/plasma). Heterogeneity is also in the types of interventions in the 23 studies that included different kinds of diets, SCFAs as capsules/infusion, or gastric bypass surgery. We were unable to perform sub-analyses such as understanding the effects of SCFA dosage or SCFA delivery route on fasting insulin and fasting glucose, due to fewer number of studies. It is also notable that majority of the studies in this analysis were conducted in three continents (Europe, the USA, and Australia), which may contribute to the risk of bias in the recruitment of individuals with a Caucasian background. However, the ethnicity of participants was not mentioned in all the studies. We recognize the differences in sensitivities of the methods used for SCFA and insulin/glucose measurement. Also, we conducted a literature search which was limited to papers published in English only, thus 169 non-English papers were excluded.

### Strengths

Despite the limitations, this is the first systematic review to summarise and provide a meta-analysis of clinical studies with the potential of increasing SCFAs. Our meta-analysis captured all studies that reported SCFA measurements and provided actual values of fasting insulin and glucose in humans. This approach was chosen as the focus of our systematic review and meta-analysis was to understand the metabolic benefits of elevated SCFA following clinical interventions.

## Conclusion

To our knowledge, this is the first meta-analysis that demonstrates a significant lowering in fasting insulin when SCFA concentrations were confirmed to have increased. Whilst such beneficial effect of SCFAs on fasting insulin has been confirmed, several factors need to be evaluated before optimal implementation such as the type of SCFA (acetate/propionate/butyrate/mixture), a form of SCFA capsules (sodium salts, inulin esters, or fibre-enriched diets), duration, and route of delivery (oral, or IV infusion) as well as compliance. Nonetheless, based on the overall results, direct SCFA consumption or a SCFA-enriched diet was found to have beneficial effects on fasting insulin. Therefore, SCFA interventions, or dietary guidance along with clinical monitoring and lifestyle management, could be considered a safe and novel treatment for overweight, obese, or subjects with type 2 diabetes.

## Data Availability

All data produced in the present study are available upon reasonable request to the authors.

## Acknowledgments

The authors acknowledge the infrastructure support through the Western Sydney University School of Medicine and the library services through the University of Technology Sydney.

## Author contributions

Conceptualization: AAH; Original search and analysis: NHTP; Validation: MVJ; Writing- first draft: NHTP, MVJ; Writing-review and editing: WKMW, NTN, AMS, AAH; Writing- finalisation: AAH. Revisions: All authors. AAH is the guarantor of this work and, as such, had full access to all the data in the study and takes responsibility for the integrity of the data and the accuracy of the data analysis.

## Supporting Information

PRISMA checklist, 2 supplementary tables, and 8 supplementary figures

