## Supplementary for "The role of short-chain fatty acids on insulin sensitivity: a systematic review and meta-analysis"

Web: https://www.isletbiology.info/

**Table S1: Previously published relevant meta-analyses and systematic reviews assessing effects of diet, pre-/pro-/synbiotics on insulin and glucose metabolism.**

| **Authors (PMID)** | **Title** | **Participants** | **Focus of the meta-analysis** | **Outcome/Conclusion** |
| --- | --- | --- | --- | --- |
| Kellow et al^28^ 2014 (24230488) | Metabolic benefits of dietary prebiotics in human subjects: a systematic review of randomised controlled trials | Healthy, Overweight, Obese, Diabetes | Effects of prebiotics on postprandial glucose, postprandial insulin, triglycerides, satiety | 26 randomised controlled trials were included in the meta-analysis. Dietary prebiotic reduced postprandial glucose and insulin concentrations. |
| Akbari V and Hendijani F^35^ 2016 (27864537) | Effects of probiotic supplementation in patients with type 2 diabetes: systematic review and meta-analysis | Diabetes | Effects of probiotics on fasting glucose, insulin, HbA1c, and HOMA-IR. | 13 clinical trials included in meta-analysis. Probiotic supplementation in T2D reduced fasting glucose & HbA1c. |
| Liu et al^30^ 2017 (27623982) | Effect of inulin-type fructans on blood lipid profile and glucose level: a systematic review and meta-analysis of randomized controlled trials | Healthy, Overweight, Obese, Diabetes | Effects of inulin type fructans on fasting glucose, fasting insulin, total cholesterol, HDL-c, and LDL-c. | 20 randomised clinical trials were included in the meta-analysis. The utilization of inulin-type fructans improved glucose control only in T2D subgroup. |
| Marventano et al^29^ 2017 (28753929) | Whole Grain Intake and Glycaemic Control in Healthy Subjects: A Systematic Review and Meta-Analysis of Randomized Controlled Trials | Healthy | Effects of wholegrains on fasting glucose, fasting insulin, glucose iAUC, insulin iAUC, maximal glucose and insulin responses, and HOMA-IR | 14 randomised controlled trials were included in the meta-analysis. Whole grains consumption improved postprandial glucose and insulin homeostasis in healthy subjects. |
| Wang et al^36^ 2017 (28599375) | Multiple effects of probiotics on different types of diabetes: a systematic review and meta-analysis of randomized, placebo-controlled trials | Overweight, Obese, Diabetes | Effects of probiotics on glucose, insulin, and HbA1c. | 18 randomized, placebo-controlled studies were included in the meta-analysis. Probiotics may offer benefits on glucose, insulin and HbA1c in T2D patients. |
| Tabrizi et al^34^ 2018 (28677046) | The Effects of Synbiotic Supplementation on Glucose Metabolism and Lipid Profiles in Patients with Diabetes: a Systematic Review and Meta-Analysis of Randomized Controlled Trials | Diabetes | Effects of synbiotics on fasting glucose, insulin, HOMA-IR, HOMA-β, QUICKI, cholesterol, triglycerides, LDL-, HDL-, and VLDL-cholesterol. | 7 randomised clinical trials were included in the meta-analysis. Synbiotic supplementation may improve glucose metabolism and lipid profiles. |
| Wang et al^31^ 2019 (31168050) | Effects of the resistant starch on glucose, insulin, insulin resistance, and lipid parameters in overweight or obese adults: a systematic review and meta-analysis | Healthy, Diabetes | Effects of resistant starch on fasting glucose, fasting insulin, HOMA-S%, HOMA-B%, HOMA-IR, HbA1c, total cholesterol, LDL-c, HDL-c, and triglycerides. | 13 case-controlled studies were included in the meta-analysis. Resistant starch improved fasting glucose, insulin, insulin resistance and sensitivity in individuals with diabetes. |
| Rao et al^32^ 2019 (31534973) | Effect of Inulin-Type Carbohydrates on Insulin Resistance in Patients with Type 2 Diabetes and Obesity: A Systematic Review and Meta-Analysis | Overweight, Obese, Diabetes | Effects of inulin-type carbohydrates on BMI, fasting glucose, fasting insulin, HbA1c, HOMA-IR, and QUICKI. | 25 studies were included in the meta-analysis. Supplementation of inulin-type carbohydrates can ameliorate insulin resistance, especially in obese with diabetes patients. |
| Ojo et al^33^ 2020 (33113929) | The Role of Dietary Fibre in Modulating Gut Microbiota Dysbiosis in Patients with Type 2 Diabetes: A Systematic Review and Meta-Analysis of Randomised Controlled Trials | Diabetes | Effects of dietary fibre on Bifidobacteria, total SCFAs, and fasting glucose. | 9 randomised controlled trials were included in the meta-analysis. Dietary fibre intervention improved HbA1c but not fasting glucose or HOMA-IR. |

Table S1: This table provides the summary for previously published systematic reviews and meta-analyses focused on different dietary interventions and their effect on fasting insulin, fasting glucose and/or HOMA-IR. The major conclusions from these systematic reviews and meta-analyses are also presented.

**Table S2: SCFA concentrations reported in different studies for the placebo and treatment groups at the end of intervention**

| **Authors** | **Concentrations of SCFAs (Mean ± SD, faecal or plasma)** | | | | | |
| --- | --- | --- | --- | --- | --- | --- |
|  | **Placebo group (Endpoint)** | | | **Treatment group (Endpoint)** | | |
|  | **Acetate** | **Propionate** | **Butyrate** | **Acetate** | **Propionate** | **Butyrate** |
| Alles et al^45^ 1999 (PMID: 9925124) | 0.11 ± 0.02 μmol/L | - | - | 0.11 ± 0.02 μmol/L | - | - |
| McIntosh (a) et al^57^ 2003 (PMID: 12663299) | 62.9 ± 17.5 μmol/g | 18.9 ± 7.9 μmol/g | 18.9 ± 9.0 μmol/g | 65.1 ± 18.0 μmol/g | 16.6 ± 4.8 μmol/g | 22.6 ± 7.9 μmol/g |
| McIntosh (b) et al^57^ 2003 (PMID: 12663299) | 62.9 ± 17.5 μmol/g | 18.9 ± 7.9 μmol/g | 18.9 ± 9.0 μmol/g | 68.5 ± 20.1 μmol/g | 18.7 ± 5.8 μmol/g | 27.8 *+* 11.6 μmol/g |
| Giacco et al^52^ 2004 | 326 ± 154 μmol/L | - | - | 302 ± 103 μmol/L | - | - |
| Maki (a) et al^56^ 2012 (PMID: 22357745) | 4.3 ± 0.66 μmol/L | 2.0 ± 0.66 μmol/L | 0.2 ± 0.2 μmol/L | 4.6 ± 0.66 μmol/L | 2.0 ± 0.66 μmol/L | 0.9 ± 0.4 μmol/L |
| Maki (b) et al^56^ 2012 (PMID: 22357745) | 4.2 ± 0.47 μmol/L | 1.8 ± 0.47 μmol/L | 0.4 ± 0.3 μmol/L | 4.4 ± 0.47 μmol/L | 1.8 ± 0.47 μmol/L | 0.1 + 0.1 μmol/L |
| Ampatzoglou et al^46^ 2015 (PMID: 25644340) | 156 ± 20 mmol/L | 46 ± 6 mmol/L | 38 ± 7 mmol/L | 199 ± 45 mmol/L | 62 ± 14 mmol/L | 54 ± 13 mmol/L |
| Sandberg et al^61^ 2016 (PMID: 6990559) | 229 ± 44 mmol/L | 3.99 ± 0.21 mmol/L | 1.74 ± 0.10 mmol/L | 251 ± 57 mmol/L | 4.38 ± 0.28 mmol/L | 2.27 ± 0.18 mmol/L |
| Chambers et al^49^ 2019 (PMID: 30971437) | 60.0 ± 28.1 μmol/L | 2.8 ± 1.0 μmol/L | 3.3 ± 0.7 μmol/L | 64.6 ± 19.1 μmol/L | 2.6 ± 1.0 μmol/L | 3.2 ± 0.3 μmol/L |
| Chambers et al^64^ (2) 2019 (PMID: 30098126) | 19.7 ± 11.1 μmol/L | 2.4 ± 0.9 μmol/L | 1.9 ± 0.6 μmol/L | 35.8 ± 16.5 μmol/L | 2.5 ± 0.9 μmol/L | 2.0 ± 0.6 μmol/L |

Table S2: This table provides the actual values of SCFAs reported in different studies assessed in this systematic review and meta-analysis. Endpoint concentrations from control and intervention groups are listed here.

**Figure S1: Forest plot of fasting insulin concentrations between control and treatment subgroup by direct or indirect SCFA administration**

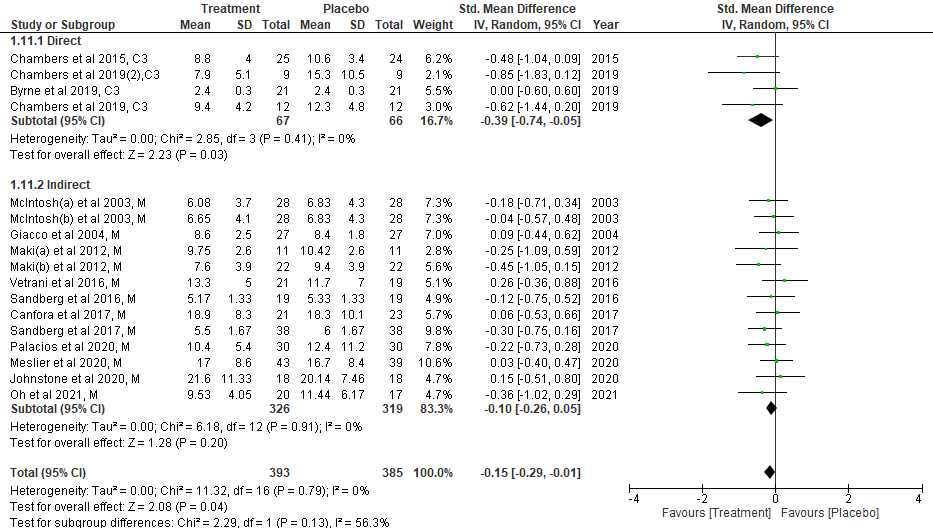
Figure S1: Fasting insulin concentrations from quantitative studies were compared between placebo and treatment groups at the endpoint (end of intervention period). Data were presented as SMD of fasting insulin (µU/mL) and was separated into two subgroups: one with direct SCFA administration (propionate via inulin-propionate ester) and another with indirect SCFA administration (different meals/diets). Type of intervention (M: meal/mixed and C3: propionate) is noted at the end of each study. df: degrees of freedom, IV: inverse variance, CI: confidence interval.

**Figure S2: Forest plot of fasting insulin concentrations between control and treatment subgroup by direct/indirect SCFA administration and SCFA increase/no increase**

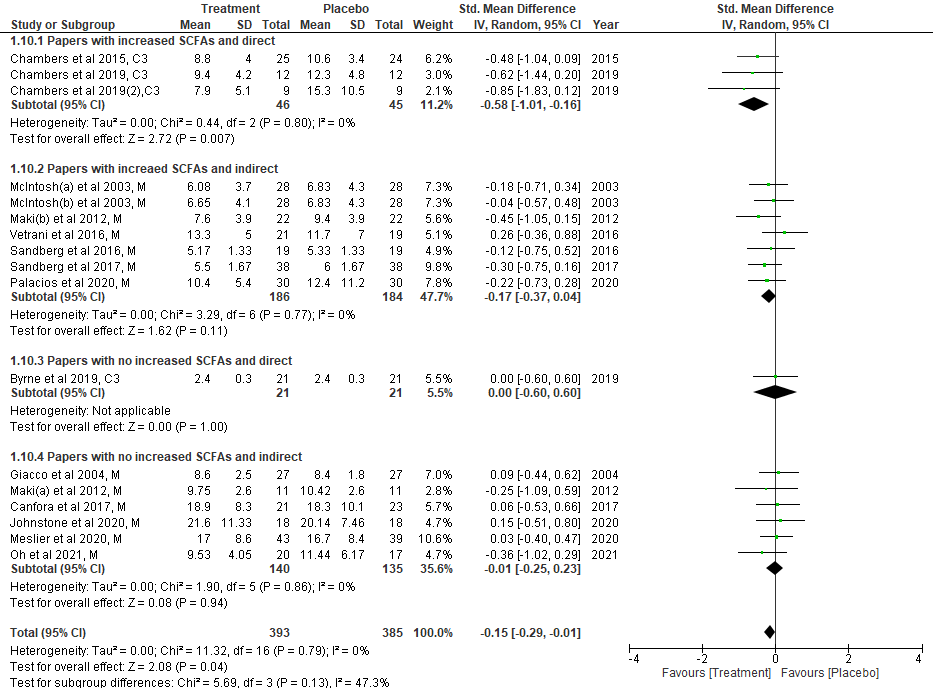

Figure S2: Fasting insulin concentrations from quantitative studies were compared between placebo and treatment groups at the endpoint (end of intervention period). Data were presented as SMD of fasting insulin (µU/mL) and was separated into four subgroups: (i) with direct SCFA administration (propionate via inulin-propionate ester) and evidence of increase in SCFAs; (ii) with indirect SCFA administration (different meals/diets) and evidence of increase in SCFAs; (iii) with direct SCFA administration (propionate via inulin-propionate ester) and no change in SCFAs; (iv) with indirect SCFA administration (different meals/diets) and no change in SCFAs. Type of intervention (M: meal/mixed and C3: propionate) is noted at the end of each study. df: degrees of freedom, IV: inverse variance, CI: confidence interval.

**Figure S3: Forest plot of fasting insulin concentrations between control and treatment subgroup by plasma/faecal SCFA measurement and SCFA increase/no increase**

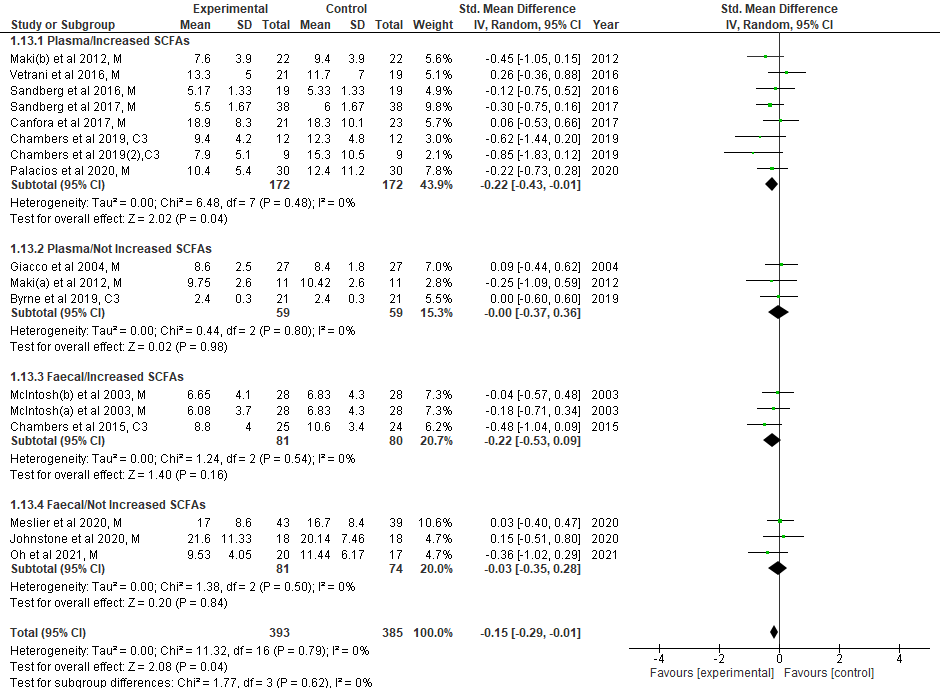

Figure S3: Fasting insulin concentrations from quantitative studies were compared between placebo and treatment groups at the endpoint (end of intervention period). Data were presented as SMD of fasting insulin (µU/mL) and was separated into four subgroups: (i) plasma SCFA measurements and evidence of increase in SCFAs; (ii) plasma SCFA measurements and no evidence of increase in SCFAs; (iii) faecal SCFA measurements and evidence of increase in SCFAs; (iv) with faecal SCFA measurements and no evidence of increase in SCFAs. Type of intervention (M: meal/mixed and C3: propionate) is noted at the end of each study. df: degrees of freedom, IV: inverse variance, CI: confidence interval.

**Figure S4: Correlation matrix between changes in SCFA, fasting insulin and fasting glucose concentrations.**

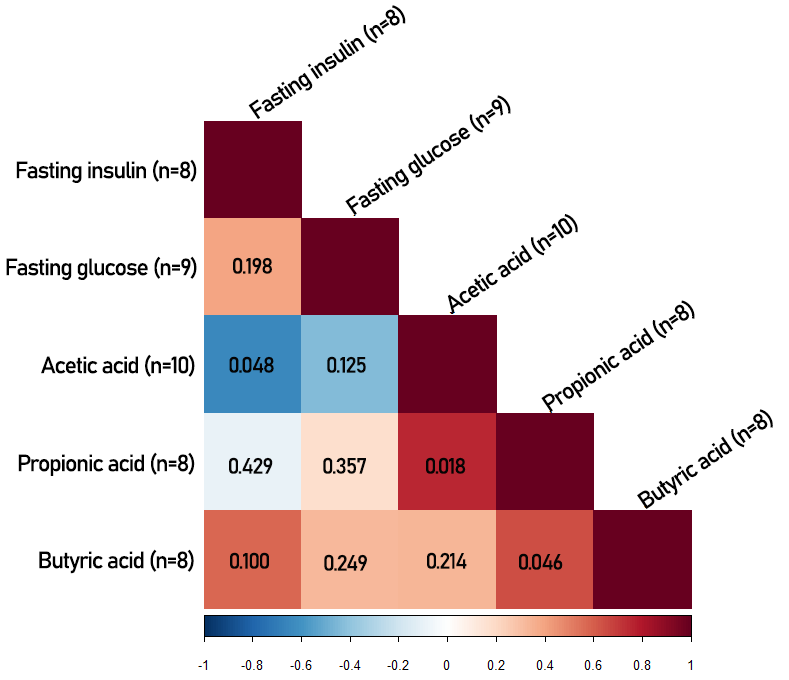

Figure S4: Correlation analysis was performed between SMD values of SCFAs, fasting insulin and fasting glucose. Eight out of 23 studies reported SCFA values for control and intervention groups at the endpoint (refer to Table S2). Spearman correlation coefficient (r) is calculated for each comparison and is presented by the colour ranging from shades of blue (negative correlation) to shades of red (positive correlation). Colour scale and corresponding r values are presented at the bottom of the matrix. One-tailed p-value for each correlation is provided as a number in respective square. Only acetate and fasting insulin demonstrated significant correlation. Number of studies included for each variable are presented in parentheses (n=8-10).

**Figure S5: Forest plot of fasting insulin concentrations between baseline and endpoint in placebo group**

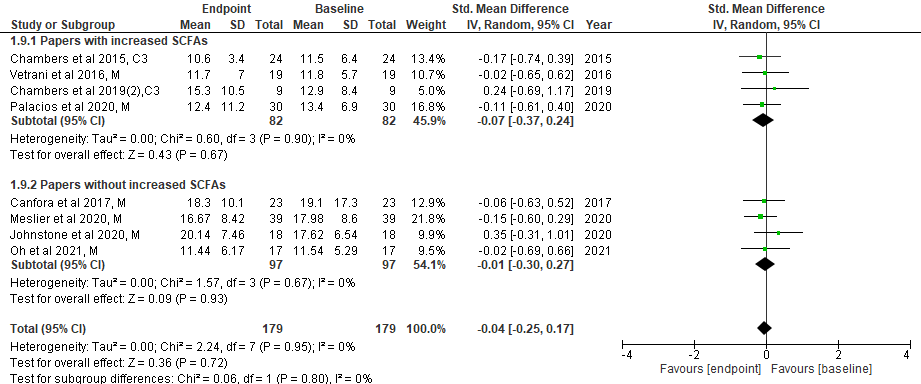

Figure S5: Fasting insulin concentrations at baseline and endpoint (end of intervention) from each study were compared in the placebo group. Data were presented as SMD of fasting insulin (µU/mL) and was separated into two subgroups: one with evidence of increase in SCFAs and another without any change in SCFAs. Type of intervention (M: meal/mixed and C3: propionate) is noted at the end of each study. df: degrees of freedom, IV: inverse variance, CI: confidence interval.

**Figure S6: Forest plot of fasting glucose concentrations between baseline and endpoint in placebo group**

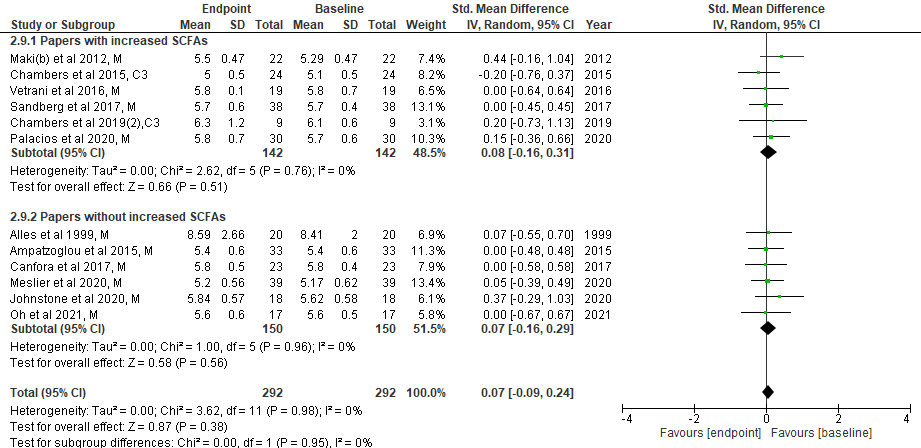

Figure S6: Fasting glucose concentrations at baseline and endpoint (end of intervention) from each study were compared in the placebo group. Data were presented as SMD of fasting insulin (mmol/L) and was separated into two subgroups: one with evidence of increase in SCFAs and another without any change in SCFAs. Type of intervention (M: meal/mixed and C3: propionate) is noted at the end of each study. df: degrees of freedom, IV: inverse variance, CI: confidence interval.

**Figure S7: Forest plot of HOMA-IR between baseline and endpoint in placebo group**

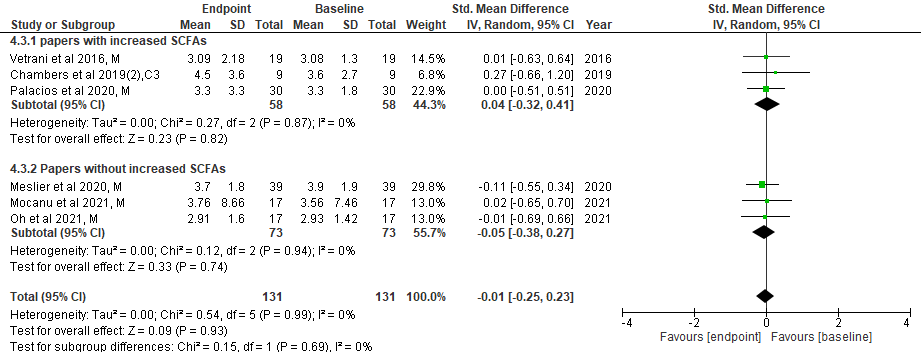

Figure S7: HOMA-IR values at baseline and endpoint (end of intervention) from available studies were compared in the placebo group. Data were presented as SMD of HOMA-IR and was separated into two subgroups: one with evidence of increase in SCFAs and another without any change in SCFAs. Type of intervention (M: meal/mixed and C3: propionate) is noted at the end of each study. df: degrees of freedom, IV: inverse variance, CI: confidence interval.

**Figure S8: Risk of bias analysis**

**A**

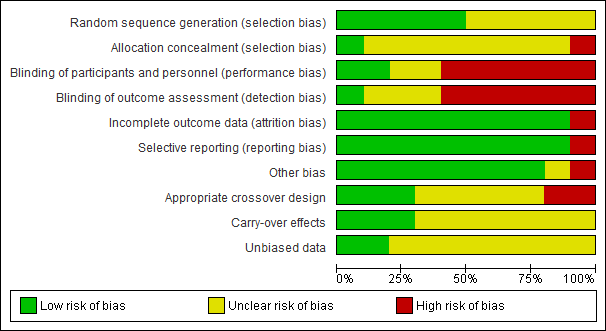

**B**

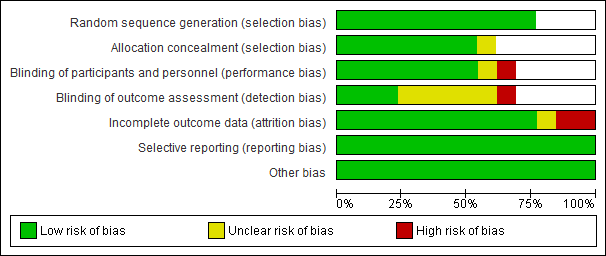

Figure S8: Risk of bias assessment of 23 papers included in the meta-analysis. Ten out of the 23 papers were crossover studies (A) and the remaining 8 papers were parallel-arm designs and 5 were treatment-alone (B). Treatment-only studies did not qualify for selection bias, performance bias and detection bias, thereby leaving them out from those risk analyses (denoted by white area in B).
